# Accuracy of online symptom checkers and the potential impact on service utilisation

**DOI:** 10.1101/2020.07.07.20147975

**Authors:** Adam Ceney, Stephanie Tolond, Andrzej Glowinski, Ben Marks, Simon Swift, Tom Palser

## Abstract

**Objectives:** The aims of this study are firstly to investigate the diagnostic and triage performance of symptom checkers, secondly to assess their potential impact on healthcare utilisation and thirdly to investigate for variation in performance between systems.

**Setting:** Publicly available symptom checkers

**Participants:** Publicly available symptom-checkers were identified. A standardised set of 50 clinical vignettes was developed and systematically run through each system by a non-clinical researcher.

**Primary and secondary outcome measures:** System accuracy was assessed by measuring the percentage of times the correct diagnosis was a) listed first, b) within the top five diagnoses listed and c) listed at all. The safety of the disposition advice was assessed by comparing it with national guidelines for each vignette.

**Results:** Twelve tools were identified and included. Mean diagnostic accuracy of the systems was poor, with the correct diagnosis being listed first on 37.7% (Range 22.2 to 72.0%) of occasions and present in the top five diagnoses on 51.0% (Range 22.2 to 84.0%). 51.0% of systems suggested additional resource utilisation above that recommended by national guidelines (range 18.0% to 61.2%). Both diagnostic accuracy and appropriate resource recommendation varied substantially between systems.

**Conclusions:** There is wide variation in performance between available symptom checkers and overall performance is significantly below what would be accepted in any other medical field, though some do achieve a good level of accuracy and safety of disposition. External validation and regulation are urgently required to ensure these public facing tools are safe.

**Strengths and Limitations:**

- Data collection was undertaken by non-clinically trained staff to replicate patient behaviour and there was random sampling to test the inter-rater reliability
- Clinical vignettes were agreed by a clinical team consisting of a GP, a pharmacist and a hospital emergency care consultant
- Current UK guidelines were used to assess service utilisation. Where symptom checkers were developed outside of the UK the disposition advice may be unlikely to be aligned due to different jurisdictions
- This research was a limited indirect study on the variety of terms and language patients might use in their interactions with these systems
- There was no assessment of how a clinician would diagnose and triage a patient presenting with the vignette symptoms

## Main text

### Introduction

In 2013, the World Health Organisation reported that the world had a 7% shortage of healthcare professionals of all disciplines which is predicted to grow to 13% by 2030. Primary care is particularly^1^ affected, with severe shortages being reported across the world^2^. For example, the United States alone is predicted to have a shortfall of between 7,300 and 43,100 primary care physicians by 2030. In the United Kingdom, a joint report by the Health Foundation, King’s Fund and Nuffield Trust in March 2019 suggested that there were 6% fewer GP’s practicing in September 2018 than in September 2015. This report further predicted there would be a shortfall of 11,500 general practitioners by 2028/29 which could become a threat to the very sustainability of primary care^3^.

This shortage has several consequences, including reduced access to health care, rising healthcare inequity, longer waiting times^2^ and increased use of emergency services.^4-5^ As a result therefore, there has been increasing interest in harnessing novel technologies, principally Artificial Intelligence (AI) techniques such as machine learning and deep learning, to reduce clinician workload, manage finite resources and help patients access the most appropriate care pathway more rapidly.

One manifestation of these new technologies is Computerised Diagnostic Decision Support (CDDS) programmes. Two broad types exist. The first are professional decision aids, most of which help stratify a patient’s risk of a particular condition or outcome and thereby help a medical professional decide what management plan they should institute. The second are “symptom checkers” which are targeted primarily at patients. Over the last decade web and app-based symptom checkers and symptom assessment tools have proliferated both in terms of geographical reach and in capability, with multiple tools now in existence, some of which claim to use AI algorithms and chatbot functionality. Their first aim is to provide information to a patient that will help them to determine the likely cause of their symptoms. The second is to provide a triage based on the symptoms presented and to advise an individual whether they should seek medical advice, and if so at what level i.e. hospital, general practice, self-care at home, and the level of urgency that is required.

The current available symptom checkers require the user to respond to a series of questions and provide a diagnosis and advice on appropriate next step. There is variability between the symptom checkers in the way individuals are asked to interact and there are different approaches to displaying endpoints for diagnostic and triage advice.

One potential benefit of such systems is the ability to identify life-threatening issues, for example stroke or heart failure, and advise the patient to seek emergency care. They also have the potential to identify less severe conditions that could be self-managed at home, thus providing reassurance to an individual by providing them with accurate information and reducing the impact on the demand for healthcare services by decreasing the likelihood individuals will access care when they can self-manage.

However, these benefits can also have the opposite impact if a symptom checking system calculates a diagnosis that is not appropriate and suggests that the individual does not seek care when they should. This could lead to false reassurance and potential harm. Individuals who could self-manage but are advised to seek care could suffer harm from unnecessary anxiety as well as creating extra demand and costs on already scarce healthcare resources, particularly in primary care.

Several previous studies^6-7^ into the effectiveness of algorithmic performance have found deficiencies in the diagnostic capabilities and a cautious approach to triage. However, only one in 2015 (Semigran et al^8^) examined multiple presentations and conditions; the others focussed on single condition studies such as those examining system performance for cervical myelopathy, inflammatory arthritis, HIV / Hepatitis C and ophthalmic conditions^9-12^. Given the refinement of existing models and the new entrants into the market since the 2015 study, the current clinical performance of these systems remains unknown.

The aim of this study therefore is to investigate the accuracy of current online symptom checkers, by assessing the diagnostic and triage performance. Secondly we assess the potential impact of the triage advice on health and care service utilisation.

## Method

### Search strategy for symptom checkers

During January 2020 the research team undertook a process to identify online symptom checkers for inclusion within the study. The approach identified tools and applications that use chatbots or algorithms to assess an individual’s symptoms and provide them with clinical advice in terms of the likely diagnosis and the suggested actions the patient should take.

In order to recreate the routine of a potential user, a systematic search was undertaken across the most popular platforms from which these products would likely be accessed. The scope of the platforms ranged from Google on the web, Google Play for Android and App store for Apple. The search terms were designed to capture a broad range of symptom checkers encapsulating the changing technologies and terminologies used by providers of these services. The search terms used can be found in supplementary table 1.

**Table 1.**
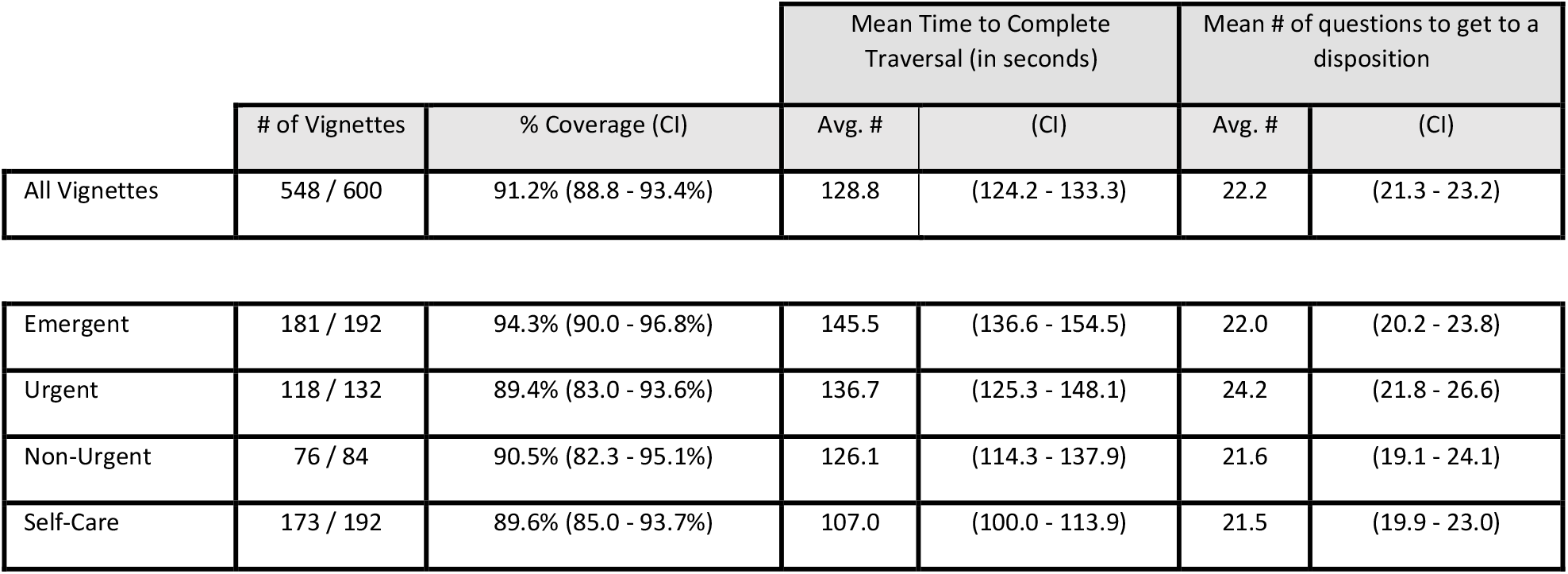
Vignette coverage and mean time to complete / questions asked.

In order to use the most popular symptoms checkers for this study, the scope of the search was limited to the first three pages of the google search and the first 25 apps on Google Play and the App Store. This is again trying to emulate “real-life” and was based on a previous study demonstrating that 97.8% of searches reached no further than the third page^13^. This generated a comprehensive list of 38 tools for potential inclusion within the study. Tools and applications were then assessed by the research team and excluded if they met any of the following exclusion criteria: (list of symptom checkers excluded and the specific exclusion reasons can be found in supplementary table 3)

**Table 2.** Accuracy of diagnostic capability first, within the top five, within a list.

|  | # of Vignettes | Accuracy of First Diagnosis |  | Diagnosis within the top 5 |  | Diagnosis found |  |
| --- | --- | --- | --- | --- | --- | --- | --- |
|  |  | # of Vignettes | % Coverage (CI) | # of Vignettes | % Coverage (CI) | # of Vignettes | % Coverage (CI) |
| All Vignettes | 548 / 600 | 206 / 548 | 37.7% (33.6 - 41.7%) | 279 / 548 | 51.0% (46.7 - 55.1%) | 297 / 548 | 54.2% (50.0 - 58.3%) |
| Emergent | 181 / 192 | 56 / 181 | 30.9% (24.7 - 38.0%) | 78 / 181 | 43.1% (36.1 - 50.4%) | 84 / 181 | 46.4% (39.3 - 53.7%) |
| Urgent | 118 / 132 | 40 / 118 | 33.9% (26.0 - 42.8%) | 59 / 118 | 50.0% (41.1 - 58.9%) | 63 / 118 | 53.4% (44.4 - 62.1%) |
| Non-Urgent | 76 / 84 | 29 / 76 | 38.2% (28.1 - 49.4%) | 40 / 76 | 52.6% (41.6 - 63.5%) | 42 / 76 | 55.3% (44.1 - 65.9%) |
| Self-Care | 173 / 192 | 81 / 173 | 47.1% (39.6 - 54.2%) | 102 / 173 | 59.3% (51.5 - 66.0%) | 108 / 173 | 62.8% (55.0 - 69.3%) |
| P-Value | - | - | 0.0155 | - | 0.0288 | - | 0.0266 |

**Table 3.** Accuracy of triage capability and safety of the advised disposition.

|  | # of Vignettes | Accuracy of disposition |  | Safety of disposition (Appropriate / Higher) |  |
| --- | --- | --- | --- | --- | --- |
|  |  | # of Vignettes | % Coverage (CI) | # of Vignettes | % Coverage (CI) |
| All Vignettes | 548 / 600 | 259 / 449 | 57.7% (53.2 - 62.2%) | 371 / 449 | 82.6% (78.9- 85.9%) |
| Emergent | 181 / 192 | 106 / 149 | 71.1% (63.4 - 77.8%) | 107 / 149 | 71.8% (64.1 - 78.4%) |
| Urgent | 118 / 132 | 50 / 96 | 52.1% (42.2 - 61.8%) | 68 / 96 | 70.8% (61.1 - 79.0%) |
| Non-Urgent | 76 / 84 | 34 / 63 | 54.0% (41.8 - 65.7%) | 55 / 63 | 87.3% (76.9 - 93.4%) |
| Self-Care | 173 / 192 | 69 / 141 | 48.9% (40.8 - 57.1%) | 141 / 141 | 100.0% (97.3 - 100%) |
| P-Value | - | - | 0.0007 | - | < 0.00001 |

The symptom checker;

- was not available for individuals to access either via the web or a dedicated application within the UK
- used another chatbot or algorithm provider as the main source
- focused on single conditions i.e. diabetes
- only focussed on paediatrics
- had narrow patient interactions where the focus was solely on condition information from an alphabetical list.

Through this process 12 symptom assessment tools were selected for further analysis and evaluation of each tool with clinical vignettes. (Further details of the symptom checkers included can be found in supplementary table 2).

### Clinical vignettes

Clinical vignettes that are commonly used to assess medical professionals’ diagnostic ability and management decisions were collated. These were selected to cover both common and uncommon conditions that might present to healthcare professionals (particularly general practitioners) in their day to day case load.

To enable comparison with previous work, the 45 clinical vignettes from the BMJ study in 2015 were reviewed by the appointed clinical team. The clinical team consisted of a GP, a pharmacist and a hospital emergency care consultant to cover all knowledge bases required for the study. Alongside this, the clinical team reviewed the common presentations in primary care^14^ and revised the vignettes to increase the breadth and coverage of conditions as described below.

From the original 45 vignettes, one pertaining to the condition rocky mountain spotted fever was removed, as it was agreed that this condition is not prevalent within the UK or Western Europe. There were an additional six vignettes sourced to cover presentations of common conditions that were not adequately represented in the 2015 paper. These covered a range of further conditions, such as depression and Covid-19 which had begun to present to primary care at the time of the vignette scoping. This made a total of 50 vignettes for inclusion.

Each of the vignettes had a defined diagnosis, case study and simplified core set of symptoms that were used by the research team’s non-clinical members to enter into the symptom checkers. This was to ensure consistency in data entry during the data collection period, thereby minimising reporting bias.

To further define the use of the vignettes they were grouped into discrete advice and triage categories using the appropriate NICE clinical knowledge summaries^15^ for the presenting diagnosis. These were emergent conditions (A&E now, Ambulance, appointment or advice within 1 hour, 16), urgent conditions (Appointment or seek advice >1hr up to and including 24hrs, 11), non-urgent conditions (Wait or seek advice more than 24 hours, 7) and self-care (15).

(see the supplementary table 4 for details on source, core symptoms, and supplemental symptoms for each vignette).

**Table 4.** Additional Resource Utilisation counts and proportions.

|  | Additional Resource Utilisation |  |
| --- | --- | --- |
|  | # of Vignettes | % Coverage (CI) |
| All Vignettes | 229 / 449 | 51.0% (46.4 - 55.6%) |
| Emergent | 73 / 149 | 49.0% (41.1 - 56.9%) |
| Urgent | 53 / 96 | 55.2% (45.3 - 64.8%) |
| Non-Urgent | 30 / 63 | 57.6% (35.8 - 59.7%) |
| Self-Care | 73 / 141 | 51.8% (43.6 - 59.9%) |
| P-Value | - | 0.7429 |

### Measures for evaluation

The evaluation of each of the symptom checkers had three distinct objectives. The first two objectives were aligned to the guidance for early stage evaluations laid out by Fraser et al^16^ and allowed the research team to permit direct comparisons between symptom checkers. Each of the objectives are now described in more detail.

The first objective was to assess algorithm performance in terms of accuracy of diagnosis, safety and advice in the event of not reaching a diagnosis. Algorithm performance was assessed by running each clinical vignette through each of the symptom checkers and recording the diagnosis and triage advice provided. The metrics used to assess the accuracy of the diagnosis were whether the correct diagnosis was:

- first in the list of possible conditions
- within the top five possible conditions listed
- if it was anywhere in the list of potentials

The safety of the disposition advice was assessed against the vignette categories and NICE guidance to check for accuracy and assess whether the advice was accurate (i.e. whether a higher or lower acuity disposition should have been recommended).

The second objective was to understand the potential impact on health and care system utilisation through the accuracy of the advice given to individuals to access the appropriate care setting for their collection of symptoms. We used the average costs of accessing a care setting in the UK i.e. A&E, GP, pharmacy. Each vignette had its service cost assessed as per NICE guidance. We assigned an additional cost to vignettes where the advice was triaged higher than national guidance recommended (risk aversion) and decreased costs where a vignette was triaged lower.

#### Analysis plan

To accurately assess the two objectives, each of the clinical vignettes was entered into each symptom checker by a single non-clinical member of the research team between the dates 25/02/2020 and 13/03/2020. Each traversal was recorded digitally and observed by another team member. For assurance of accuracy when entering symptoms into the checker and collating results, a random sample of 15% of vignettes for each symptom checker was assessed by a different team member to check the consistency of results and behaviours.

Following this process, the team calculated summary statistics for diagnostic accuracy, triage advice, mean number of questions and time to complete a traversal with 95% confidence intervals based on binomial distribution using R-Studio version 3.5.1. The significance of the difference in mean rates was assessed using Chi^2^ tests and R^2^ was used to evaluate the significance of correlation.

### Patient and Public Involvement

Patients and the public were not involved in this research.

### Patient Consent

Patient consent is not required as the clinical vignettes developed for research purposes are fictional accounts of presentations related to the diagnosis in focus and are not related to any individuals.

## Results

### Sample size and coverage

The search identified twelve assessment tools which met the study criteria. They were registered to operate in the United Kingdom, United States, Germany, Spain and Poland (supplementary table 2). Seven tools provided both diagnostic and triage advice, two only provided diagnostic advice with limited triage for ‘red flag’ events, and three only provided triage advice.

Algorithmic performance was assessed on a total of 600 traversals of the clinical vignettes across the 12 tools with completion of 548 or 91% (95% CI 88.8% − 93.4%; Table 1). Of the 52 traversals that could not be completed 45 (86.5%) were due to age restrictions within the systems, six (11.5%) were due to symptoms not being covered by the systems and one (1.9%) was due to system failure.Across the symptom checkers, ten of the twelve (83.3%) asked for input of demographic information (age and sex) which was be used to support the diagnostic and triage capabilities.

### Accuracy of diagnosis

Overall, across all vignettes evaluated, the correct diagnosis was listed first in 37.7% of traversals (95% CI 33.6 − 41.7%, Table 3). Performance varied significantly (p=0.0155) across the four triage categories, with the performance improving as the urgency of the condition became less.

The correct diagnosis was listed first on 30.9% (95% CI 24.7 − 38.0%) of occasions for the emergent group, 33.9% (95% CI 26.0 − 42.8%) for the urgent group, 38.2% (95% CI 28.1 − 49.4%) for the non-urgent group and 47.1% (95% CI 39.6 − 54.2%) for the self-care group.

Performance also varied between the different systems (Figure 1 &2). The percentage of times in which the correct diagnosis was listed first ranged from 22.2% (95% CI 13.6 −35.2%) in the least accurate system to 72.0% (95% CI 52.8 − 88.0%) in the most (P = <0.0001). A positive correlation was observed (R^2^ = 0.7126, Figure 1) that the more questions asked, and the longer time taken on average during a traversal, the more accurate the diagnostic result appeared to be. For example, the most accurate system used a mean of 45.8 questions and took a mean of 217.4 seconds to traverse, whereas the least accurate used a mean of 9.5 questions and took a mean of 37.7 seconds to traverse. It was also noted that this least accurate system did not ask for demographic information. The correlation became less significant when diagnosis was within the top 5 (R^2^ = 0.3056, Figure 2).

**Figure 1.**
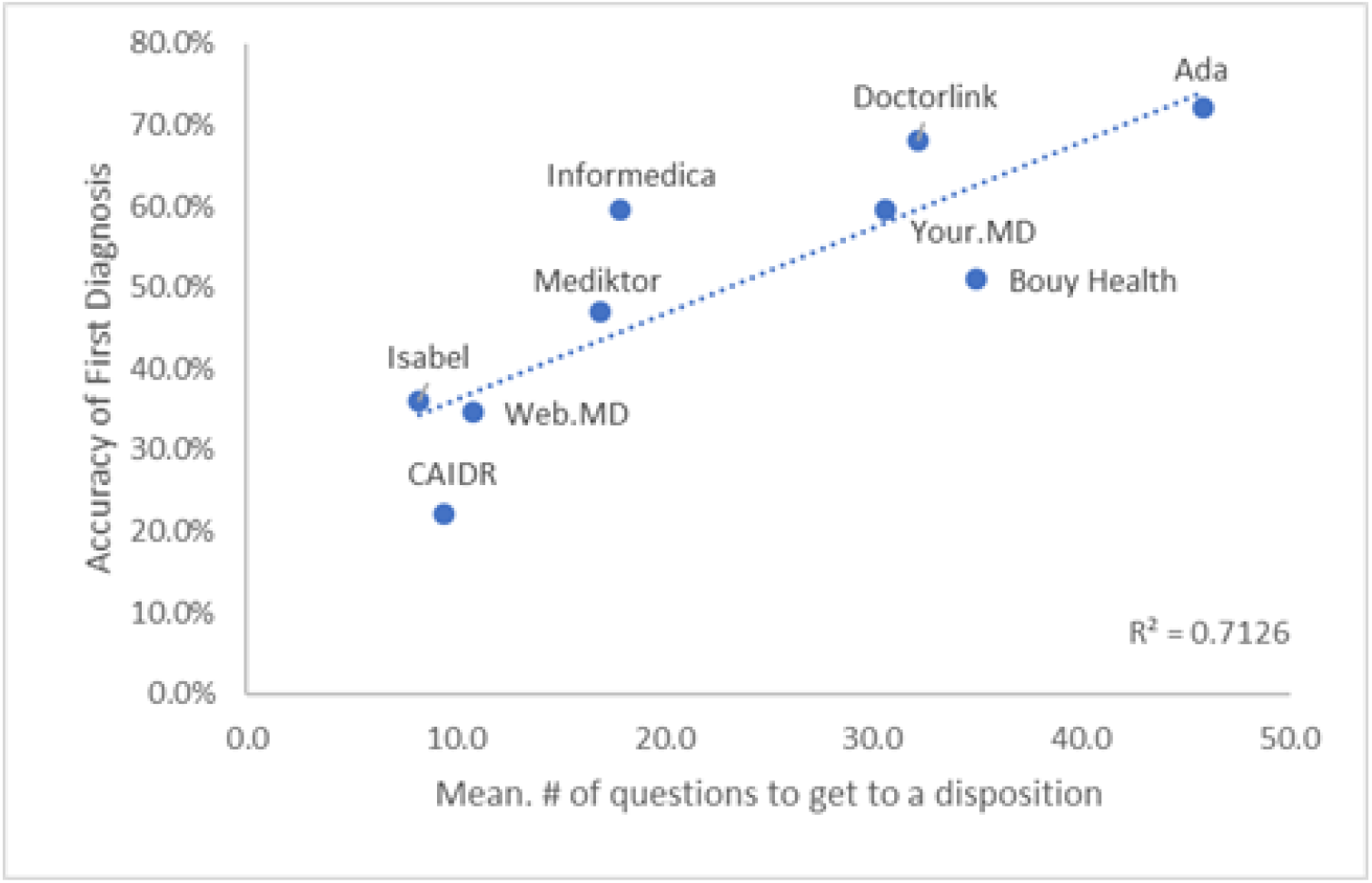
Correlation of mean number of questions to diagnostic accuracy listed first.

**Figure 2.**
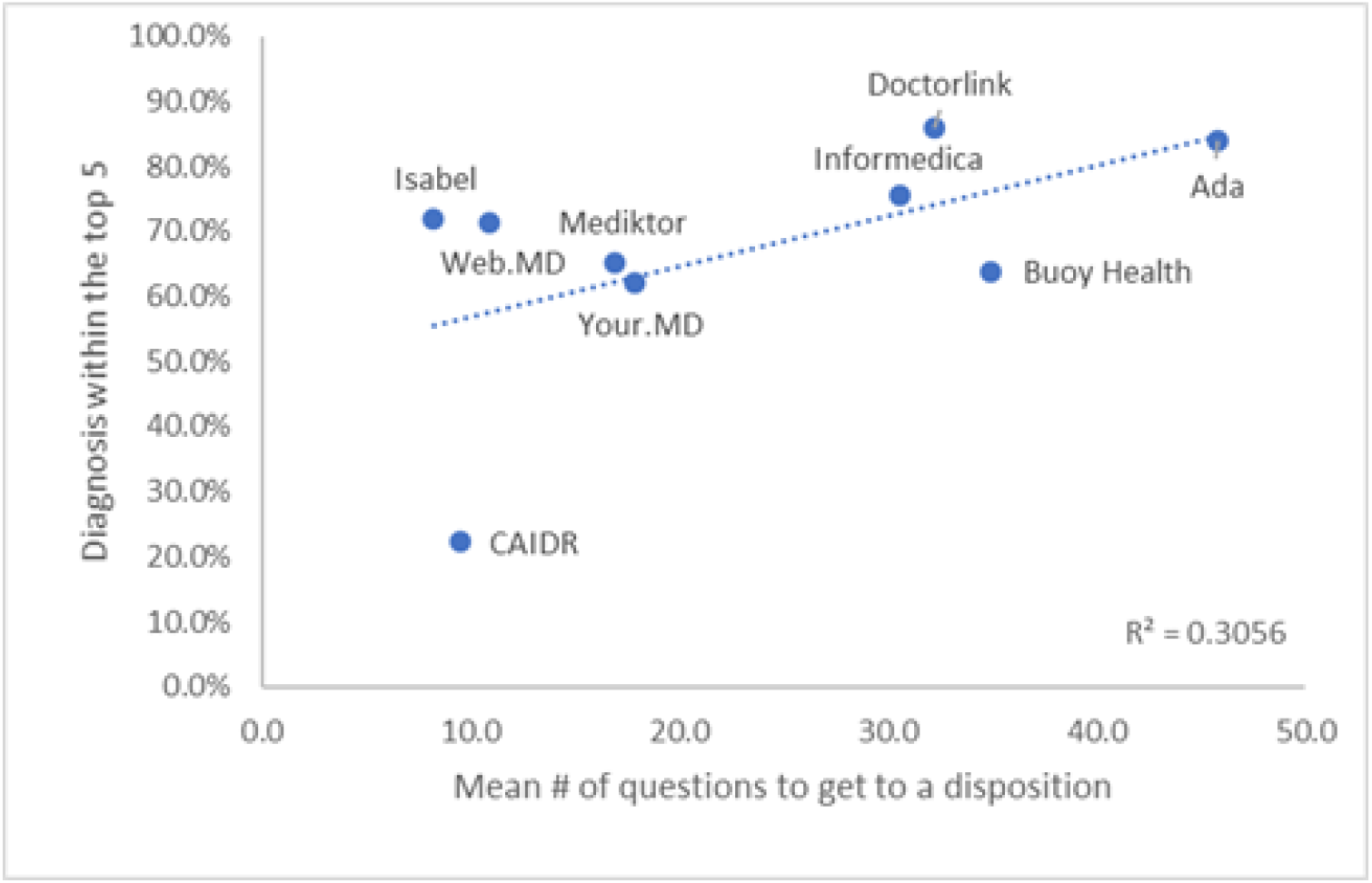
Correlation of mean number of questions to diagnostic accuracy listed in the top five.

Across all symptom checkers the correct diagnosis was listed in the top five diagnoses 51.0% (95% CI 46.7-55.1%) of the time and the likelihood of the correct diagnosis listed at all was 54.2% (95% CI 50.0 −58.3%). For both metrics, diagnostic accuracy was higher in the lowest urgency triage category (self-care) than in the highest (the emergent group) which was statistically significant with P-values for both metrics at <0.0001.

### Accuracy of triage advice

Appropriate triage advice across all clinical vignettes tested was given in 57.7% of cases (95% CI 53.2 – 62.2%;Table 3). Performance varied across the four categories with the emergent care group being statistically higher at 71.1% (95% CI 63.4 − 77.8%; p-value .000686) then the other three groupings which all performed below 50% with the urgent group at 52.1% (95% CI 42.2 − 61.8%), non-urgent group at 54.0% (95% CI 41.8 − 65.7%) and the self-care group at 48.9% (95% CI 40.8 − 57.1%).

Again there was significant variation in performance between symptom checkers with a range between 35.6% (95% CI 24.4 −49.0%) to 90.0% (95% CI 69.1% − 100%) across all vignettes tested (P = <0.0001) (Figure 3). This however there was not a significant correlation (R^2^ = 0.1344, Figure 3) with the number of questions asked to reach the triage advice. It was observed that there was a mixed approach to triage advice with some systems being specific i.e. ‘see a GP within 8hrs’ and others with a broader approach of ‘seek medical care’ which may create less certainty for individuals accessing the system.

**Figure 3.**
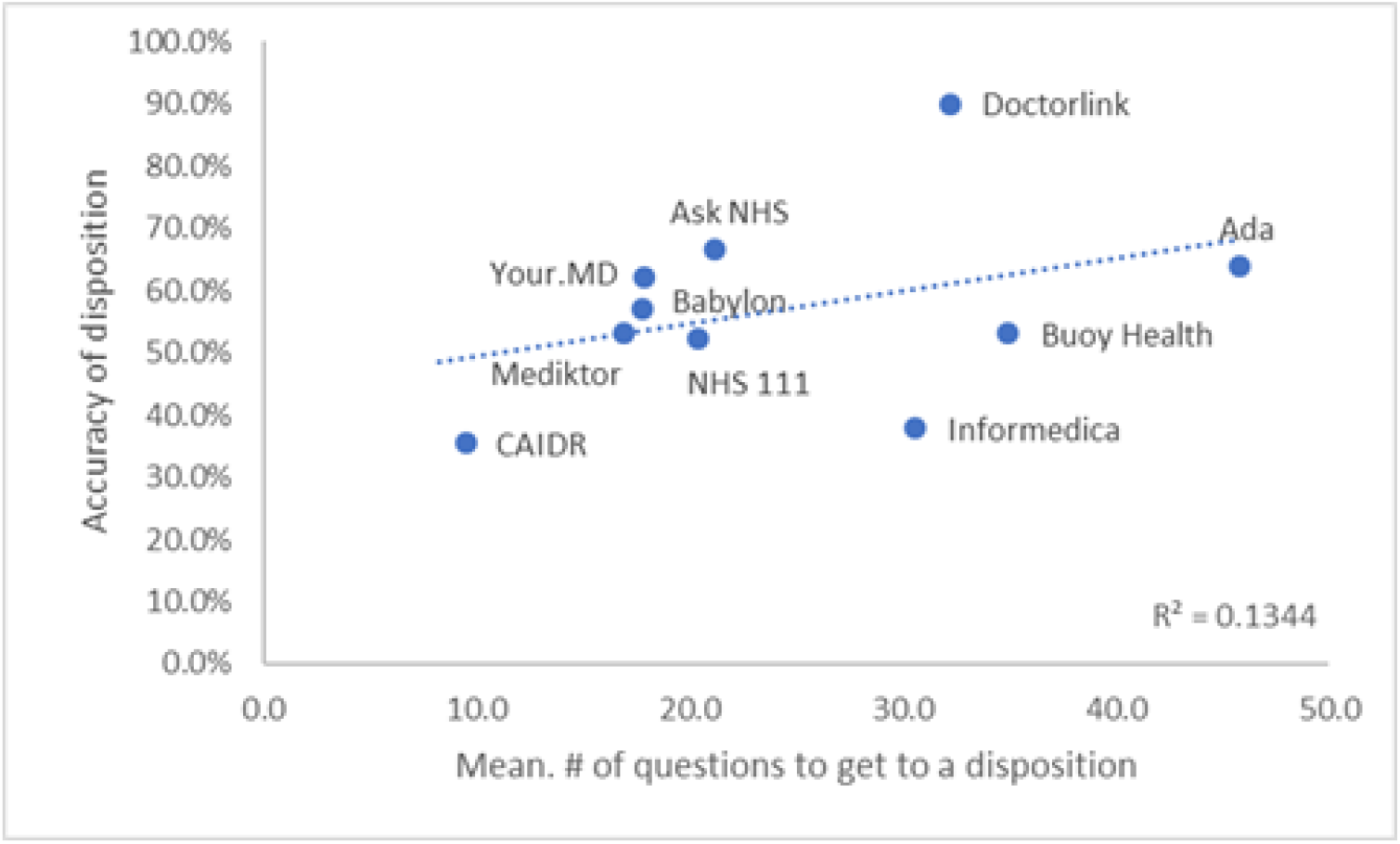
Correlation of mean number of questions to triage accuracy.

The safety of the triage advice across all clinical vignettes was appropriate in 82.6% (95% CI 78.9-85.9%) of all vignettes. For this metric, safety was higher in the lowest urgency triage category (self-care) than in the highest (the emergent group) which was inverse to the accuracy of triage where higher urgency (the emergent group) was higher than the lowest urgency self-care group (P = <0.0001) (Figure 4).

**Figure 4.**
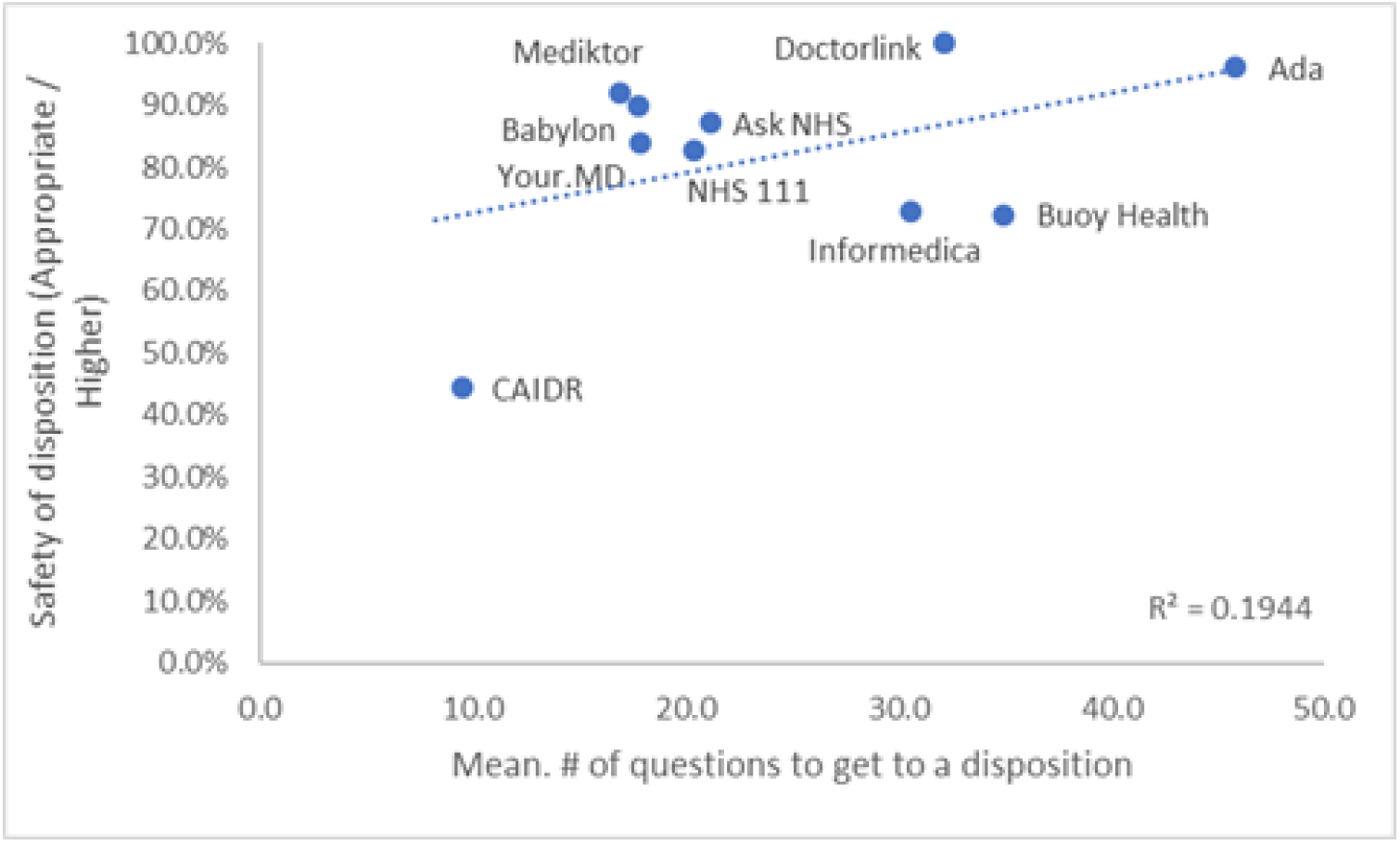
Correlation of mean number of questions to triage safety. The relationship between triage accuracy and triage safety is important and largely linear (R^2^ = 0.6074, Figure 5) with some providers of symptom checkers scoring highly on both, some symptom checkers however, are less accurate particularly with regard to accuracy of disposition. (See fig 5)

### Impact on services

Dispositions advice was compared against current NICE clinical knowledge summaries. Across all vignettes completed, 51.0% (46.4-55.6%, p-value .7429 ; Table 4) required additional resource utilisation than that recommended. Where self-care would be the most appropriate disposition according to NICE clinical knowledge summaries 51.8% (95% CI 43.6-59.9%) of completed traversals suggested a health care service was required.

There was variation in performance (Figure 6,7) across providers with additional resource utilisation ranging between 12.5% (95% CI 6.1-33.5%) for the lowest impact symptom checker and 87.5% (95% CI 52.8%-100%) for the highest (P = <0.0001). Of the total traversals where NICE clinical knowledge summaries indicate self-care, the symptom checkers advised a primary care visit in 80.8% of cases where additional resource was required.

**Figure 5.**
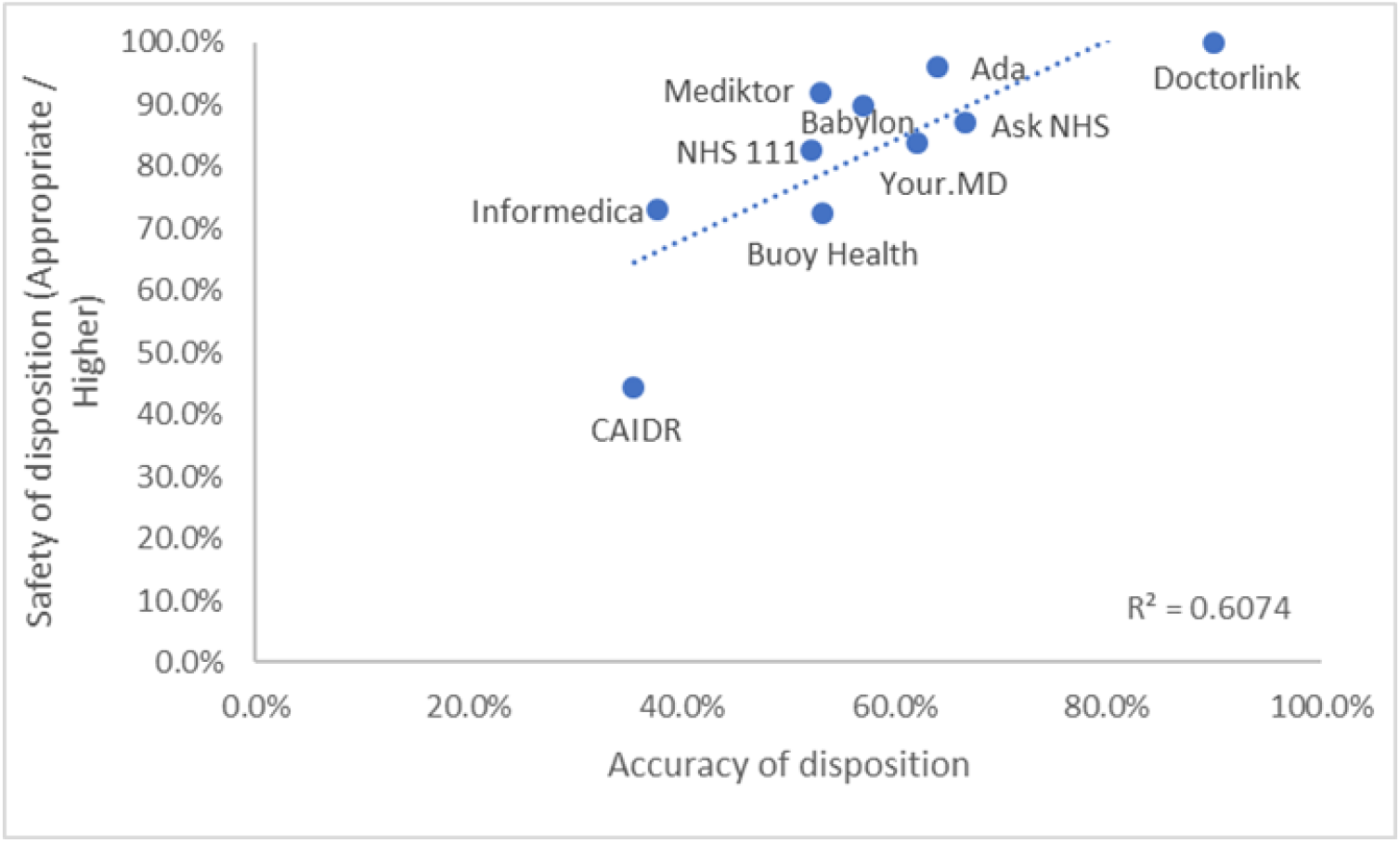
Correlation of triage safety and accuracy of disposition.

**Figure 6.**
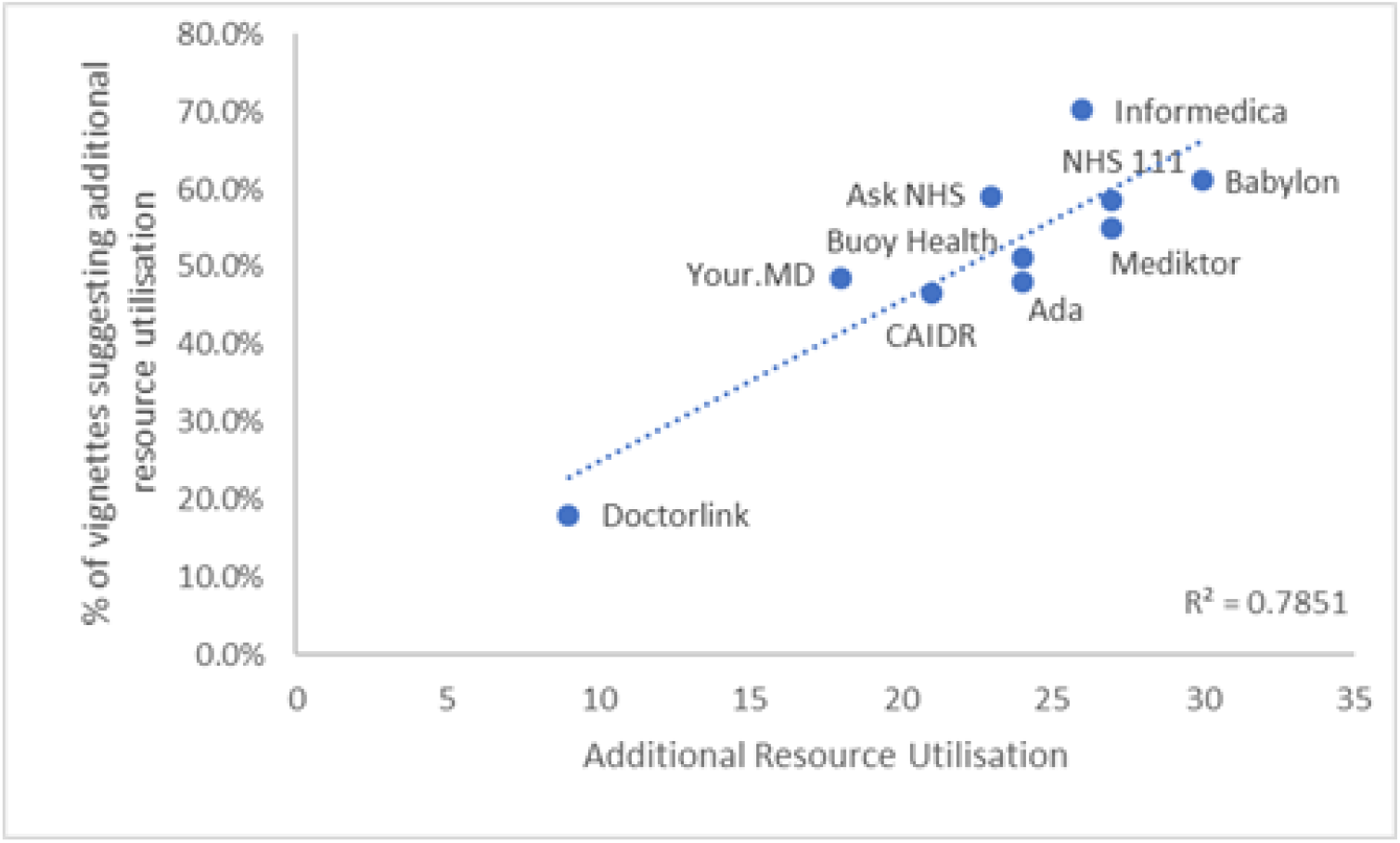
Correlation of additional resource units for all vignettes and self-care specific.

**Figure 7.**
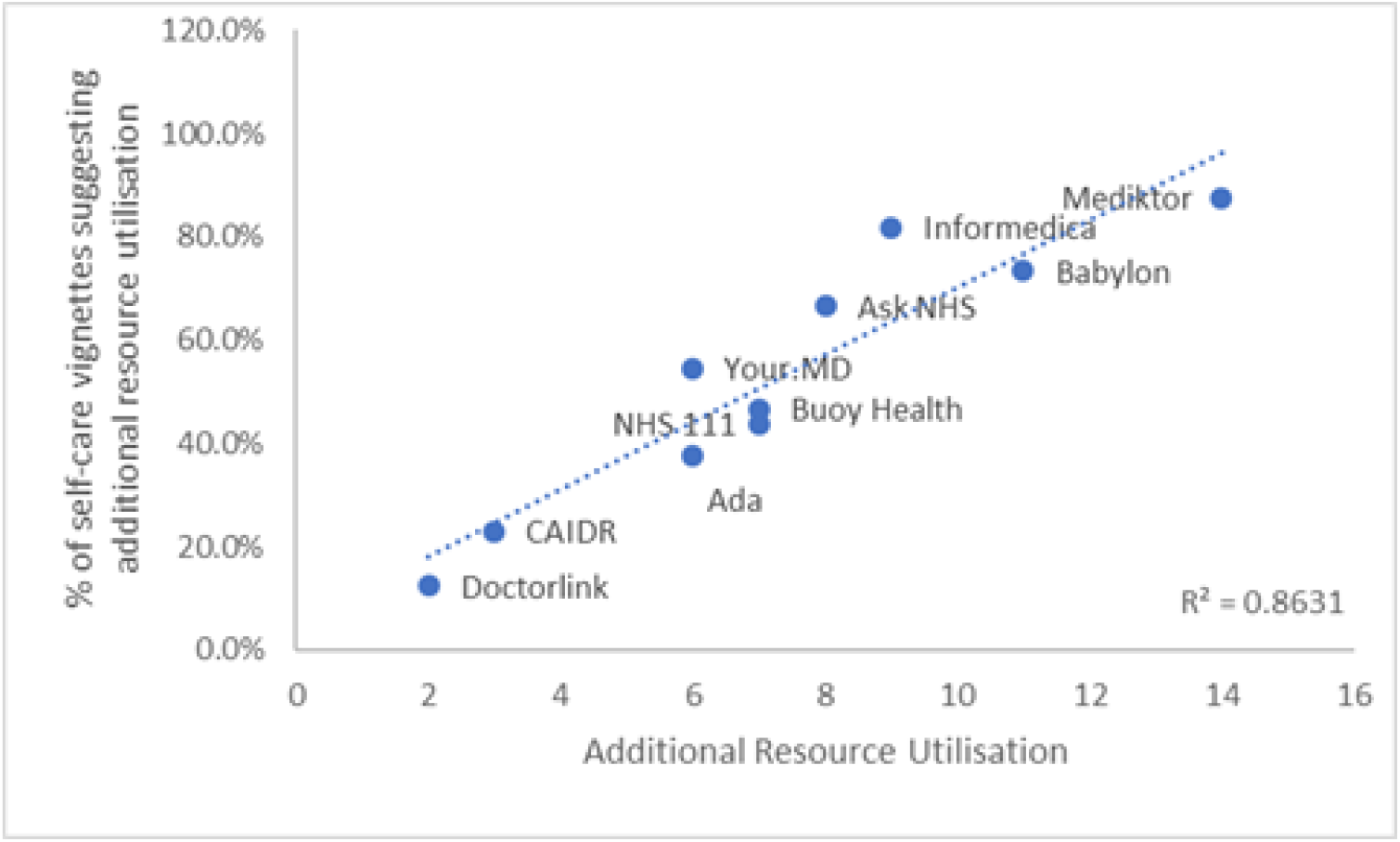
Correlation of additional resource units for self-care specific vignettes.

Across the other three categories, 49.3% of additional service utilisations suggested were for the highest cost resources of ambulance call outs or a visit to the emergency department. Primary care visits accounted for a further 39.7% with 6.8% “seeking medical advice” which would most likely be primary care and 4.1% being an urgent treatment centre.

## Discussion

### Summary

Our primary findings were that the average diagnostic accuracy of symptom checkers was poor, being correct on only just over a third of occasions (37.7%) and present in the top five diagnoses on only approximately half (51.0%) of occasions.

Secondly, the accuracy with which the appropriate level of care was recommended by the systems was low, being accurate just over half the time (57.7%), though some systems demonstrated high levels of accuracy of disposition and safety. We found little evidence that they can currently reduce healthcare resource use, as we found that that additional resource utilisation was suggested in over half of cases, with primary care resources being suggested in 80.8% of conditions that could be self-managed.

Finally, the analysis found that there was wide and significant variation in the performance of different systems, both in terms of diagnosis and in the appropriateness of the recommendation given. The percentage of times in which the correct diagnosis was listed first ranged from % (95% CI 13.6 −35.2%) in the least accurate system to 72.0% (95% CI 52.8 − 88.0%) in the most,whilst the accuracy of the care recommendation varied from 35.6% (24.4 −49.0%) to 90.0% (69.1% − 100%). Likewise systems varied in resource use, with half of the systems recommending additional healthcare service use in more than 50% of patients in whom self-care is appropriate (range 12.5% − 87.5%).

This variation in system performance is a key finding. As Fraser et al^16^ discuss in their Lancet editorial, there is currently little or no regulatory oversight of these systems (and although industry-funded, this is the first external examination of the sector in five years). Patients have no way of knowing if the system they find on a search engine is accurate or not (and many probably assume they are).

As Fraser et al comment “systems that are poorly designed or lack rigorous clinical evaluation can put patients at risk and likely increase the load on health systems”. Systems that inappropriately recommend additional health resource use are likely to put additional strain on already stretched public health systems. They may also induce unnecessary worry and anxiety for patients and induce them to pay for unnecessary private care. Our study therefore reinforces Fraser’s findings and we echo their call for urgent, external oversight of these systems.

### Comparison with existing literature

When compared with the Semigran study, the overall diagnostic capabilities of the symptom checkers assessed were similar, with the correct first diagnosis being suggested at 34% in the Semigran paper and 37% in this study. For overall triage disposition there was no difference between the results of this study and the Semigran study, which also reported 57%. There was however a statistically significant difference in the triage accuracy for the self-care grouping (33% vs 48.9%) which suggests that the symptom checkers assessed have developed in their capability to provide support and information to individuals to within this category. Notably, some systems did have higher performance indicating that some systems have used advancing technology to improve their accuracy but importantly this is not uniform across the sector.

This study has gone further than the Semigran study in that it has attempted to contextualise the risk averse behaviour that has previously been seen in this and other studies^9-12^ .This study demonstrates that individuals are being recommended to access services that their symptoms do not warrant by many of the symptom checkers assessed, potentially putting additional pressure on resources and adding undue worry on individuals that they must seek medical care. Although perhaps not surprising given the increasingly litigious nature of healthcare on both sides of the Atlantic, this is a notable concern. One of the oft-quoted potential advantages of these systems is that they can reduce demand on increasingly over-stretched health systems. This study indicates that many systems do not currently function well enough for this potential advantage to be realised.

One issue is that there may be confusion about the actual purpose of symptom checkers, whether that be to reduce service load, improve access to care, help with patients’ information needs or a mix of the three. It is likely that one use is for rapid and accessible guidance, with the average time to complete a traversal being just over two minutes and given they are available 24 hours, 7 days a week. Patients who experienced delays in accessing care due to difficulties obtaining an appointment or conflicts between surgery opening times and their work commitments were more than twice as likely to use on-line information services^18^. One thing that does appear to be clear is that they are popular with patients. One example study examining a specific tool found over 80% of patients perceived it to be useful and over 90% would use it again^17^. With the increasing shortfall in workforce and resources, the use of on-line services such as symptom checkers is only likely to increase, so these shortfalls in performance are a major concern.

### Strengths and limitations

Our study had several strengths and limitations. Firstly non-clinically trained staff were used to undertake the data collection in order to replicate the conditions of an untrained individual accessing each system. As some of the vignettes contained specific clinical language, the simplified language was enhanced and any reference to clinical tests that an individual presenting with the same symptoms would potentially use were removed. This however is only in the view of the individuals involved in the study and, whilst random sampling, to test inter-rater reliability, was undertaken on 15% of vignettes for each symptom checker, this study is an indirect assessment of the variety of terms and language an actual patient may use in their interactions with these tools. Whilst clinicians used the NICE clinical knowledge summaries and agreed on the dispositions, this was done by committee and does not assess how well a clinician would diagnose or triage an individual presenting in a real-life clinical setting. It was also observed that tools developed outside of the UK do not always have recommended dispositions that align with the NICE guidelines and may be appropriate for the jurisdiction within which they have been developed.

Finally, this was an observational study that allowed for the analysis of data entered on an individual’s symptoms or on behalf of someone to compare machine recommendations with current clinical guidance. Further evaluations on available symptom checkers should include formal field trials, including randomised controlled trials for impact evaluation^16^ to assess if use of the symptom checker leads to better health outcomes than usual care in clinical settings. We fully agree that that systems should be regularly tested once they are made publicly available to ensure performance remains in line with disease and symptom prevalence, and as people become more aware and better able to make decisions about their health and wellbeing. This aligns with the suggestions by Fraser et al as part of their study. ^16^

### Implications for research and/or practice

There has not been a peer reviewed study into the efficacy of online symptom assessment tools since the BMJ paper in 2015. Five years have passed, and technology has been enhanced with the increasing maturity of natural language processing (NLP) and artificial intelligence (AI). There are now many new entrants into the market, and so it is important to continually re-evaluate the systems’ clinical performance with a wide range of clinical conditions to understand not only performance, but the impact the decision points may have on the healthcare system.

Despite the development in algorithmic technologies there still remain many deficiencies in the diagnostic and triage capabilities and wide variability in performance between systems, to an extent that would be unacceptable in any other form of medical device, instrument or therapy. The advice given by poor quality symptom checkers may encourage individuals to access unnecessary care, which puts pressure on increasingly stretched healthcare systems, particularly primary care (as demonstrated in this study). To actively take part in their health and wellbeing individuals require access to high quality, evidenced-based information. Symptom checkers have significant potential to help in the ability to provide this but only if they are robust, transparent and accurate. They should not be viewed as a replacement for traditional clinician triage but as one part of a toolkit that could in time become first line support for advice and guidance.

If their potential is to be realised and for them not to have the converse effect to their intent, with negative impacts and an increase rather than a reduction in resource use, it is important that further controlled trials on how individuals receive and interpret information presented to them are performed. Most importantly it is essential that these symptom checkers should be routinely audited, with the results made public to ensure that they remain current to clinical guidelines and their accuracy and the standard of the services they provide is transparent to the public and as high as any other area of medical practice.

### Contributorship statement

SS, AC, AG and BM conceived and designed the study. Data analysis was performed by AC, ST and TP;

AC and TP prepared the manuscript. All named authors read, revised and approved the final manuscript. AC is guarantor. No other individual satisfying these criteria has been excluded from authorship.

This article is respectfully submitted as an original article.

## Data Availability

The Corresponding Author has the right to grant on behalf of all authors and does grant on behalf of
all authors, a worldwide licence to the Publishers and its licensees in perpetuity, in all forms, formats
and media (whether known now or created in the future), to i) publish, reproduce, distribute, display
and store the Contribution, ii) translate the Contribution into other languages, create adaptations,
reprints, include within collections and create summaries, extracts and/or, abstracts of the
Contribution, iii) create any other derivative work(s) based on the Contribution, iv) to exploit all
subsidiary rights in the Contribution, v) the inclusion of electronic links from the Contribution to
third party material where-ever it may be located; and, vi) licence any third party to do any or all of
the above.

## Competing interests

All authors have completed the Unified Competing Interest form at www.icmje.org/coi_disclosure.pdf (available on request from the corresponding author) and declare:

Doctorlink employees had sight of the manuscript and were able to make comments on it, but all decisions were made independently by the authorship team.

A copy of the draft manuscripts that they viewed, with all suggestions, was attached with the submitted article for transparency purposes.

## Data sharing statement

The Corresponding Author has the right to grant on behalf of all authors and does grant on behalf of all authors, a worldwide licence to the Publishers and its licensees in perpetuity, in all forms, formats and media (whether known now or created in the future), to i) publish, reproduce, distribute, display and store the Contribution, ii) translate the Contribution into other languages, create adaptations,reprints, include within collections and create summaries, extracts and/or, abstracts of the Contribution, iii) create any other derivative work(s) based on the Contribution, iv) to exploit all subsidiary rights in the Contribution, v) the inclusion of electronic links from the Contribution to third party material where-ever it may be located; and, vi) licence any third party to do any or all of the above.

